# Discriminating Inflammation from Malignancy with Short-Dynamic Patlak Parametric 18F-FDG PET/CT

**DOI:** 10.1101/2025.09.30.25336991

**Authors:** Jelena Jandric, Lorenzo Leonardi, Rossella Barisonzi, Roberta Zanca, Carlo Vallone, Marcello Rodari, Laura Evangelista, Alessia Artesani

## Abstract

**Aim/Introduction:** Differentiating malignant from inflammatory uptake on ^18^F-FDG PET/CT remains a major diagnostic challenge, as standardized uptake value (SUV) lacks specificity. Dynamic acquisitions with Patlak analysis can separate metabolized from unmetabolized tracer, potentially improving discrimination. We evaluated whether short-duration dynamic FDG PET/CT with Patlak parametric imaging provides complementary information beyond SUV for distinguishing malignancy from inflammation.

**Materials and Methods:** Twenty-seven patients undergoing oncologic PET/CT (breast, lung, or gastrointestinal cancer) were included, yielding 96 lesions (69 malignant, 27 inflammatory). Short dynamic acquisitions (20 min) were motion-corrected and analysed to generate influx rate (*K*_*i*_) and distribution volume (*V*_*d*_) maps. Lesions were segmented on SUV images (40% SUVmax), and radiomic features were extracted from SUV, *K*_*i*_, and *V*_*d*_ maps. Exploratory data analysis, linear modelling, and dimensionality reduction assessed separability. A Random Forest classifier was trained with crossvalidation, integrating Synthetic Minority Oversampling (SMOTE) to address class imbalance. An independent validation cohort of 15 lesions (13 inflammatory, 2 malignant) was tested.

**Results:** Malignant lesions showed higher SUVmean (5.8 vs. 2.8 g/ml) and *K*_*i*_ (1.95 vs. 0.75 ml/min/100ml), whereas inflammatory lesions demonstrated higher *V*_*d*_ (44.7 vs. 35.1%). No single feature provided reliable thresholds. Logistic regression achieved 89% accuracy but suffered from quasi-separation, confirming limited linear discriminability. Random Forest classification yielded robust performance (cross-validated AUC-ROC 0.876; AUC-PR 0.948). With G-mean thresholding, inflammation was detected with high recall (0.93) but recall for malignancy was lower (0.74). Feature importance highlighted SUV and *K*_*i*_ variance, as well as *K*_*i*_/ *V*_*d*_ ratios, as strongest predictors. In the external validation set, accuracy reached 0.80, with inflammation reliably identified (precision 0.85, recall 0.85).

**Conclusion:** Short dynamic Patlak imaging combined with machine learning improves the characterization of malignant versus inflammatory uptake beyond SUV alone. By decomposing FDG uptake into metabolized (*K*_*i*_) and unmetabolized (*V*_*d*_) fractions, this approach provides physiologically meaningful separation of tracer behaviour. While sensitivity for malignancy requires further optimization, our findings establish a reproducible framework for future more extensive research on clinical interpretation of parametric imaging in oncologic PET.

## I. INTRODUCTION

[^18^F] Fluorodeoxyglucose (FDG) Positron Emission Tomography (PET) coupled to Computed Tomography (CT) is a cornerstone of oncologic imaging because many malignancies exhibit elevated glucose metabolism [1]. However, FDG is not tumour-specific, and some inflammatory cells can also overexpress glucose transporters and hexokinase activity, leading to avid FDG uptake at sites of infection or inflammation [2]. Because of this lack of specificity, inflammatory processes may mimic malignancy on FDG PET, posing a diagnostic dilemma. FDG-PET studies noted incidental accumulations in granulomatous or infectious foci during routine cancer scans [3], underlining that benign processes may appear with elevated uptake on PET.

In practice, nuclear medicine physicians rely on uptake patterns, intensity, and relationships to areas of physiological distribution, as well as clinical context to suggest inflammation versus tumour. Common inflammatory patterns on PET/CT include diffuse uptake (as in colitis), spread or segmental uptake (as in vasculitis) or hilar nodal uptake (as in sarcoidosis), whereas malignancies often appear as more focal masses [4][5]. Nevertheless, such patterns frequently overlap. Moreover, current EANM/SNMMI guidelines for FDG PET in inflammation note that SUV thresholds are not validated in infection or inflammation [2] and should be used with caution. For example, one study advised SUVmax*>*3 to improve specificity in spinal infection [6], but such cut-offs remain empirical. In fact, relatively high SUV may be seen in inflammation, as well as relatively low SUV may be seen in malignant lesions. False positives are well-documented – for example, post-surgical inflammatory reactions or uptake around implants (vascular grafts, prostheses) can mimic tumour [7]. In short, no standardized PET criteria exist to distinguish inflammation from cancer, and interpreting equivocal FDG foci often depends on reader experience, additional imaging, or biopsy.

To address this challenge, dynamic PET/CT with kinetic modelling has emerged as a promising tool. In dynamic scanning, tracer activity is recorded over time, and graphical Patlak analysis is applied to compute voxel-wise parameters. This yields quantitative images of the FDG influx rate constant (*K*_*i*_, related to the Patlak slope) –proportional to the metabolic rate of FDG trapping – and the vascular distribution volume (*V*_*d*_, related to the Patlak intercept) representing un-metabolized tracer in blood. These parametric maps separate the metabolic (irreversible) component of uptake from blood-pool effects [8]. In several studies, Patlak-derived *K*_*i*_ images frequently show consistently higher lesion-to-background contrast than conventional SUV images [9][10][11]. In some cases, combining SUV with Patlak-derived metrics appears to improve diagnostic accuracy. Zhang et al. reported that incorporating both metabolic-rate (*K*_*i*_) and distribution-volume (*V*_*d*_) images increased the specificity and accuracy of lesion characterization [12]. Recently, simplified protocols like short dynamic acquisitions have been shown to yield reliable *K*_*i*_ estimates while minimizing scan time [13]. These developments suggest that kinetic parameters could in principle enhance specificity for tumour versus inflammation, although formal guidelines on their use have not yet been established.

Early clinical studies support this idea. Skawran et al. performed dynamic whole-body FDG PET in patients with both cancer and inflammatory lesions; they found that the Patlak *K*_*i*_,max was significantly higher in malignant targets than in infectious/inflammatory ones (mean 3.0 vs. 2.0 ml/min/100ml, p=0.002) [14]. However, the overlap in *K*_*i*_ and in lesion-to-background ratios (LBR) was large – e.g. LBR(*K*_*i*_) was 5.0 vs 4.4 (p=0.05) for cancer vs inflammation [14]. Similarly, a total-body dynamic PET study using an ultra-long scanner found that malignant lymph nodes had much higher *K*_*i*_ (2.6 ml/min/100ml) than benign nodes (0.66 ml/min/100ml) (p*<*0.001), whereas *V*_*d*_ did not differ significantly between benign and malignant nodes [15]. Nonetheless, no clear indication, consensus criteria or cutoffs have been defined. In other words, dynamic Patlak imaging remains a research tool rather than a standardized clinical procedure.

Building on these findings, we sought to investigate whether short-duration dynamic Patlak FDG PET/CT can improve the distinction between malignant and inflammatory uptake. Our study focuses on patients with breast, lung, or gastrointestinal cancer who underwent PET/CT for staging or restaging and in whom FDG-avid lesions were identified. Inflammatory findings were also included when present as comorbid conditions such as tendinitis, fractures, or muscle inflammation. We hypothesize that Patlak-derived kinetic parameters provide complementary information beyond conventional SUV, particularly in equivocal cases. Specifically, we expect that *K*_*i*_ (reflecting irreversible metabolic trapping) and *V*_*d*_ (reflecting vascular distribution) differ systematically between malignant and inflammatory processes. By extracting radiomic features from both static SUV images and Patlak-derived parametric maps, we aim to quantitatively compare malignant lesions with benign inflammatory findings and determine whether exists a linear or non-linear combination able to estimate the risk of malignant versus inflammatory lesion.

## II. MATERIALS AND METHODS

### A. Population cohort

The cohort for this observational study consisted of 27 patients undergoing short dynamic FDG PET/CT scans for the evaluation of suspected or confirmed breast, lung or gastrointestinal cancer, with inflammation also detected in some cases. Table I reports indications for age, sex, injected dose and disease type in our cohort.

**Table I.** Patient characteristics. Values are given as median (range) for continuous variables and number (percentage) for categorical variables.

|  |  |
| --- | --- |
| **Age** (year) | 60 (range: 23-84) |
| **Sex**, n(%) |  |
| male | 11 (41%) |
| female | 16 (59%) |
| **Injected Dose** (MBq) | 293 (range: 197 – 362) |
| **Disease**, n(%) |  |
| Gastrointestinal Cancer | 6 (22%) |
| Breast Cancer | 7 (26%) |
| Lung Cancer | 5 (19%) |
| Other solid tumours | 3 (11%) |
| Without evidence of malignancy | 6 (22%) |

### B. PET/CT Acquisition

A short-dynamic whole-body (dWB) PET/CT protocol from SAFOV scanner (Biograph Vision 600, Siemens Healthineers) was performed. Patients received a weight-based FDG injection (4.8 MBq/kg), followed by a 50-minute uptake period. A dWB PET/CT scan was then acquired using continuous bed motion (20 minutes total, 4 × 5-min passes). A low-dose CT scan was acquired first for attenuation correction and anatomical localization. PET images were reconstructed using an OSEM algorithm (4 iterations, 5 subsets), incorporating time-of-flight and resolution modelling. Corrections were applied for attenuation, scatter, random coincidences, dead-time, and radioactive decay. No postreconstruction filtering was used. The resulting images comprised 335 planes (440 × 400 voxels) with a voxel size of 1.65 × 1.65 × 3 mm^3^. All PET data were fully anonymized prior to analysis. An image-derived arterial input function (AIF) mask was generated by manually drawing a circular region of interest (approximately 15 mm in diameter) in the descending aorta.

Before running the Patlak graphical analysis, the dynamic PET frames were corrected for motion, in agreement with previously published work [16]. Specifically, we used the open-source Phyton-based FALCON software (version V2) [17][18], specifically tailored for correcting motion among PET frames. In this study, we performed deformable registration with 50 × 50 × 50 iterations at each level of the multi-scale registration. The registration started from the 1st frame and used the last frame as reference, as this frame is used to generate the diagnostic SUV image and further guarantee correct co-registration between parametric results and SUV images. Details on the FALCON software in correcting both rigid and nonlinear motion artifacts in dynamic whole-body images are demonstrated in a previously paper published by the group working at the ENHANCE initiative [19].

Patlak parametric images were generated using an indirect reconstruction workflow, as motion correction required data export and frame-wise processing. In this study, we used an open-source Python pipeline (`pyPatlak`) to compute voxel-wise *K*_*i*_ and *V*_*d*_ maps from motion-corrected dynamic PET frames. The code was validated against commercial direct Patlak analysis and source code is available on GitHub [20][21].

To summarize, the general workflow included the following steps:

- **Data export and motion-corrected frames**: PET DICOM data were exported, anonymized and corrected for motion artifact. This step ensures the spatial alignment across frames.
- **Input function normalization**: A centre-specific population-based input function (PBIF), previously validated in our institution [22], was used as reference curve. The PBIF was scaled to patientspecific AIF by extracting a volume-of-interest (VOI) in the descending aorta from the reconstructed PET data.
- **Voxel-wise Patlak analysis**: Using pyPatlak, voxel-wise linear regression was to derive *K*_*i*_ and *V*_*d*_ maps.

### C. Lesion delineation

A board-certified nuclear medicine physician segmented malignant and inflammatory lesions on static SUV PET images. Areas of increased FDG uptake relative to the background, corresponding to either malignancy or inflammation, were identified and segmented using a 40% SUVmax threshold in the Siemens Healthineers syngo.via workstation. This threshold defined the volume of interest (VOI) for each lesion. The resulting VOI masks were exported and applied to both SUV and Patlak parametric images, enabling extraction of quantitative metrics including mean, maximum, and variance values, as well as first-order radiomic features (Skewness, Kurtosis, Entropy). Data were stored in CSV format and subsequently analysed with Patlak graphical method.

### D. Statistical Analysis

#### Data preparation

Data preprocessing included the removal of samples according to predefined exclusion criteria. To avoid significant partial-volume effects, lesions with diameter smaller than 10 mm were excluded. Intestinal parametric maps were also omitted from analysis, as the motion-correction algorithm could not fully compensate for peristaltic motion, potentially compromising parameter accuracy. Ultimately, liver lesions were excluded due to physiological uptake heterogeneity, and significantly high value of parameter *V*_*d*_ in this region. The final dataset comprised 96 lesions, classified as malignant (69) or inflammatory (27), and 15 lesions classified as ‘unknown’ as used for the validation test and comprises of both malignant (2) and inflammatory (13) lesions. Each lesion was characterized by three quantitative imaging variables (SUV, *K*_*i*_ and *V*_*d*_) and six first-order radiomic features (mean, maximum, variance, skewness, kurtosis, entropy).

#### Exploratory Data and Linear Separability Analysis

All statistical analyses were conducted in Python (v. 3.10.3). A preliminary exploratory data analysis was performed to characterize the dataset and assess relationships between malignant and inflammatory lesions. Class distribution was determined, and descriptive statistics (median and interquartile range) were computed for all quantitative imaging and radiomic features. Normality was assessed using the Shapiro–Wilk test, and differences between malignant and inflammatory lesions were evaluated using the Mann–Whitney U test. Feature interdependence was examined through Pearson correlation analysis to identify potential multicollinearity. Linear separability was then assessed by testing whether the two classes could be distinguished using a simple linear model. Univariate logistic regression was performed for each feature, and variables with p *<* 0.10 were included in a multivariate model. In the multivariate logistic regression, all tests were two-sided, with statistical significance set at p *<* 0.05. This step established a baseline for evaluating the performance of linear models in differentiating lesion types. To further explore class separability, dimensionality reduction techniques were applied. Principal component analysis (PCA) and t-distributed stochastic neighbour embedding (t-SNE) were used to visualize the distribution of lesions in a two-dimensional feature space. Prior to dimensionality reduction, standard scaler was used to remove intensity differences amongst parameters.

#### Random Forest Classifier

A machine learning model was developed and validated to differentiate malignant from inflammatory lesions with non-linear model. Two feature sets were evaluated: (1) the “All Features” set, comprising all 18 available radiomic metrics, and (2) the “SUV-only” set, restricted to 7 features derived exclusively from PET images. A Random Forest Classifier (RFC) was selected as the classification algorithm. All features were normalized using a `MinMaxScaler` within the model pipeline to mitigate the impact of differing feature scales and units. Model performance was assessed using 5-fold stratified cross-validation, ensuring preservation of the original class distribution across folds and providing an unbiased estimate of performance on unseen data. Hyperparameter tuning was performed via ‘GridSearchCV’, optimizing parameters including the number of estimators (`n_estimators`), maximum tree depth (`max_depth`), minimum samples required to split a node (`min_samples_split`), and minimum samples per leaf (`min_samples_leaf`). The model was evaluated based on the Area Under the Receiver Operating Characteristic Curve (AUC-ROC). Given the class imbalance in the dataset (28% inflammatory vs. 72% malignant lesions), G-mean threshold optimization was applied to select a classification threshold that balanced sensitivity and specificity across both classes. The optimal threshold was defined as the point maximizing the geometric mean (G-mean) of sensitivity and specificity, penalizing solutions that performed poorly for either class.

To further address class imbalance, the Synthetic Minority Over-sampling Technique (SMOTE) was integrated directly into the cross-validation pipeline. SMOTE generates synthetic samples for the minority class (inflammatory lesions) by interpolating between each minority instance and its k-nearest neighbours. Crucially, SMOTE was applied only to the training data within each cross-validation fold, preventing data leakage and ensuring that model performance was evaluated exclusively on untouched test sets.

## III. RESULTS

### A. Results from Exploratory Data and Linear Separability Analysis

A total of 96 samples were included in the analysis following data cleaning, with a class imbalance (72% malignant vs. 28% inflammatory). Descriptive statistics (median and interquartile range [IQR]) for all radiomic features are provided in Table S1 (Supplementary Materials). Here, we focus on differences in mean values between the two classes (Table II). Malignant lesions generally exhibited higher SUV mean values (5.8 g/ml, IQR = 4.9) compared with inflammatory lesions (2.8 g/ml, IQR = 0.9). However, some malignant tumours displayed SUV values as low as those observed in inflammation. A similar pattern was found for parametric images: the average *K*_*i*_ was 1.95 ml/min/100 ml for malignancies and 0.75 ml/min/100 ml for inflammatory lesions. While malignant lesions tended to show higher SUV and *K*_*i*_ values, these differences did not reach statistical significance. Importantly, not all lesions with SUV greater than 3 g/ml could be confidently classified as malignant (Figure 1-Left). By contrast, *V*_*d*_ demonstrated an opposite trend, with higher mean values in inflammatory lesions compared to malignant ones. Nonetheless, no clear threshold could be defined to reliably separate the two classes (Figure 1-Right). These results highlight the diagnostic challenge of differentiating malignant from inflammatory lesions using SUV, *K*_*i*_, or *V*_*d*_ in isolation. They highlight the need for a more comprehensive, multifactorial approach to improve lesion characterization.

**Table II.** Mean values and 95% confidence intervals (CI) of SUV, *K*_*i*_, and *V*_*d*_ for malignant and inflammatory lesions. SUV = standardized uptake value; *K*_*i*_ = influx rate constant; *V*_*d*_ = distribution volume.

| Class | SUV (g/ml) | $K_i$ (ml/min/100ml) | $V_d$ (%) |
| --- | --- | --- | --- |
| Inflammation | 2.8 (2.4, 3.1) | 0.75 (0.59, 0.92) | 44.7 (37.3, 52.1) |
| Malignancy | 5.8 (5.0, 6.5) | 1.95 (1.66, 2.22) | 35.1 (29.6, 40.7) |

**Figure 1.**
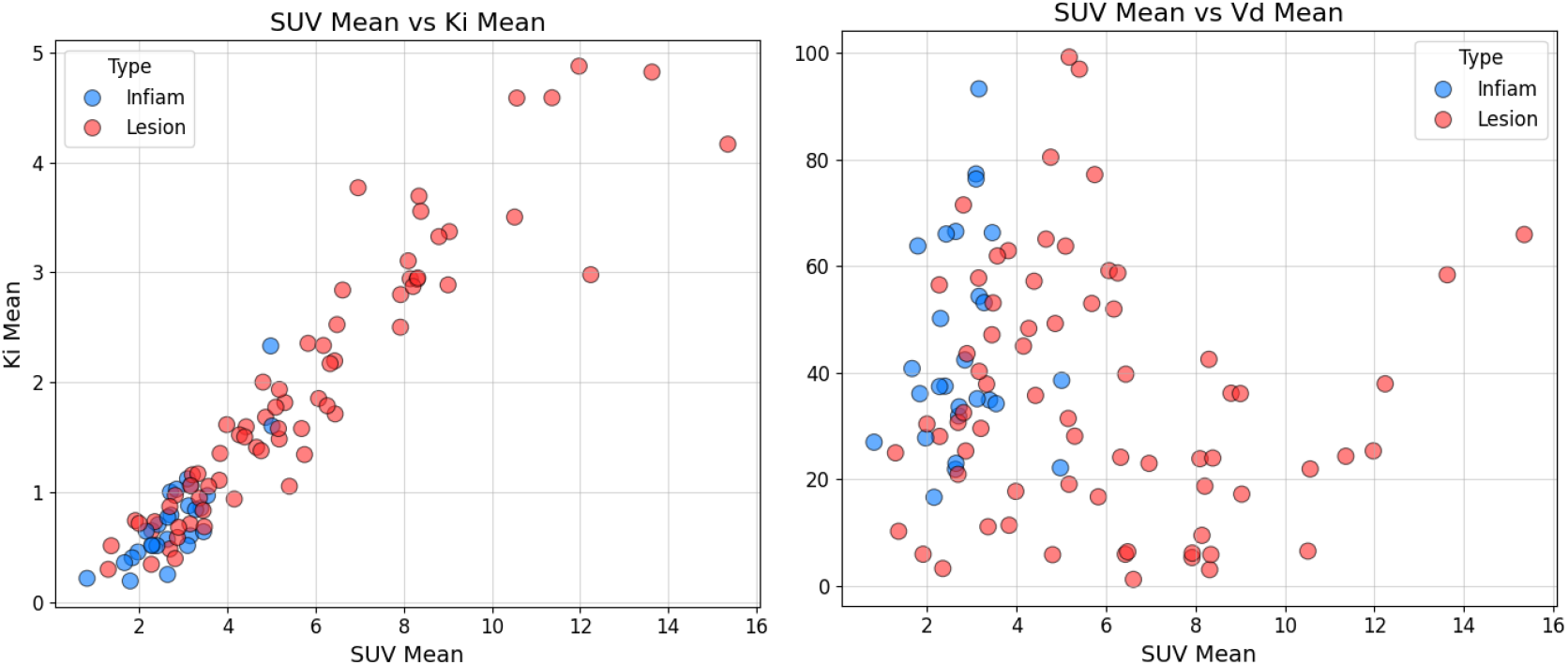
Scatterplots illustrating relationships between SUV, *K*_*i*_, and *V*_*d*_. (Left) SUV mean versus *K*_*i*_ mean, showing higher *K*_*i*_ values in malignant lesions. (Right) SUV mean versus *V*_*d*_ mean, showing higher *V*_*d*_ values in inflammatory lesions. SUV = standardized uptake value; *K*_*i*_ = influx rate constant (ml/min/100ml); *V*_*d*_ = distribution volume (%)

Correlation analysis revealed strong interdependence among features, especially within SUV- and *K*_*i*_-derived metrics (r *>* 0.70), indicating substantial multicollinearity and potential instability in linear models (Figure S1-A). Single-feature classification analysis (Figure S1-B) confirmed SUV- and *K*_*i*_-based metrics as the strongest predictors. SUV Variance yielded the highest AUC (0.854; sensitivity = 0.768, specificity = 0.889), while SUV Mean and *K*_*i*_ Mean also performed well (AUCs = 0.843 and 0.837, respectively), with very high specificity (*>*0.92) but only moderate sensitivity (0.70). *K*_*i*_ Entropy achieved a more balanced but modest performance (AUC = 0.747). By contrast, *V*_*d*_-derived metrics performed poorly, with AUCs close to chance and very low sensitivities (*<*0.20).

To establish a linear baseline, logistic regression was applied. Univariate analysis identified 13 candidate features (all SUV features except Skewness; all *K*_*i*_ features except Entropy; plus, *V*_*d*_ Mean, Skewness, and Entropy) with p *<* 0.10, which were then included in the multivariate model. Although the model converged and achieved 89% accuracy, quasi-separation was observed, suggesting that some observations were perfectly classified—a sign of potential model instability. Moreover, high p-values for individual predictors confirmed their limited independent discriminative power, indicating that lesion classification cannot be adequately captured by linear relationships.

To further examine class separability, dimensionality reduction was performed after feature standardization. PCA showed that the first two components explained 62% of the total variance, with balanced contributions across multiple features rather than dominance by any single variable. Visualization with PCA and t-SNE confirmed substantial overlap between malignant and inflammatory lesions (Figure S3), demonstrating that they are not linearly separable. Together, these findings highlight the complex, non-linear structure of the data and support the need for more robust non-linear models, such as Random Forest classifiers, for reliable lesion classification.

### B. Random Forest Classification

The initial model, trained on the original imbalanced dataset, demonstrated a robust discriminatory capability with an average AUC-ROC of 0.876, AUC-PR of 0.948. The McNemar’s test showed no statistically significant difference between the “All Features” and “SUV-only” sets (p = 0.479), although the inclusion of all radiomic features yielded a slightly higher average AUC. The optimal classification threshold, determined by G-mean maximization, was 0.768, reflecting a cautious bias toward classifying lesions as malignant. This resulted in a trade-off: high recall for inflammation but lower recall for malignancy (Table III). The skewed threshold highlights the model’s difficulty in establishing a balanced decision boundary under class imbalance.

**Table III.** Random Forest classification report (cross-validation) for all radiomic features (SUV + Parametric images) and SUV-only features. Precision, recall, F1-score, and support are reported for each class.

| Classification Report (ALL features) |  |  |  |  |
| --- | --- | --- | --- | --- |
| Class | Precision | Recall | F1-Score | Support |
| Inflammation | 0.60 | 0.93 | 0.72 | 27 |
| Malignancy | 0.96 | 0.74 | 0.84 | 69 |
| Accuracy |  |  | 0.80 | 96 |
| Classification Report (SUV features only) |  |  |  |  |
| Class | Precision | Recall | F1-Score | Support |
| Inflammation | 0.54 | 0.81 | 0.65 | 27 |
| Malignancy | 0.91 | 0.72 | 0.81 | 69 |
| Accuracy |  |  | 0.75 | 96 |

The integration of SMOTE within the cross-validation pipeline improved both model stability and interpretability. Although overall AUC gains were modest, McNemar’s test revealed a statistically significant difference between models trained with all features versus SUV-only features (p = 0.052). This occurs because SMOTE balances the dataset, reduces variance across folds, and produces more systematic prediction errors, thereby increasing the statistical power of McNemar’s test. Clinically, this indicates that comprehensive radiomic features provide meaningful predictive information beyond SUV-derived parameters, supporting their integration into diagnostic workflows. Feature importance consistently highlighted Variance, Mean, and Maximum from SUV and *K*_*i*_ metrics as strongest predictors (Table IV). Notably, ratio-based features such as maximum *K*_*i*_/*V*_*d*_ and variance of *K*_*i*_/*V*_*d*_ demonstrated strong discriminative power, with AUCs of 0.839 and 0.881, respectively. Variance of *K*_*i*_/*V*_*d*_ exhibited the highest specificity (0.963), mar*K*_*i*_ng it as a reliable indicator for excluding false positives.

**Table IV.** Feature importance analysis from Random Forest classifier with SMOTE. Optimal thresholds, sensitivity, specificity, and AUC are provided for the six most predictive features.

| Feature | SUV Mean | SUV Var. | $K_i$ Mean | $K_i$ Entropy | Max $K_i/V_d$ | Var. $K_i/V_d$ |
| --- | --- | --- | --- | --- | --- | --- |
| Optimal threshold | 3.57 | 0.70 | 1.16 | 3.62 | 0.021 | 0.0006 |
| Sensitivity (TPR) | 0.71 | 0.77 | 0.68 | 0.77 | 0.81 | 0.64 |
| Specificity (TNR) | 0.92 | 0.89 | 0.93 | 0.74 | 0.74 | 0.96 |
| AUC | 0.84 | 0.85 | 0.84 | 0.75 | 0.84 | 0.88 |

Comparing SUV, *K*_*i*_ and *V*_*d*_ provided biological insight: inflammatory lesions had limited uptake (on average below 3.5 g/ml), a more uniform distribution of unmetabolized tracer, but localized trapped FDG peaks. On the contrary, malignant lesions have medium-high uptake, more widespread irreversible FDG metabolic trapping, and highly homogenous distribution of unmetabolized tracer —consistent with altered vascular delivery and increased intracellular phosphorylation. These results support parametric imaging as a complementary diagnostic tool to SUV, enabling quantitative separation of tracer delivery and metabolism.

### C. Validation Test

To further evaluate model performance, a validation test was performed on 15 segmented lesions (13 inflammation/2 malignancy) not included in the training dataset. The same radiomic analysis was performed on these lesions and the model was asked to identify whether malignant or inflammatory. The ground truth classification was provided by the nuclear medicine physician, while the Random Forest model outputs were compared accordingly. The results are summarized in Table V. When all radiomic features were used, the model achieved an accuracy of 0.80, with 11 true negatives, 1 true positive, 2 false positives, and 1 false negative. Inflammatory lesions were recognized with high reliability (precision = 0.85, recall = 0.85), whereas malignant lesions were less consistently identified (precision = 0.33, recall = 0.50).

**Table V.** Random Forest classification report (cross-validation) for all radiomic features (SUV + Parametric images) and SUV-only features. Precision, recall, F1-score, and support are reported for each class.

| Validation Test (ALL features) |  |  |  |  |
| --- | --- | --- | --- | --- |
| Class | Precision | Recall | F1-Score | Support |
| Inflammation | 0.85 | 0.85 | 0.85 | 13 |
| Malignancy | 0.33 | 0.50 | 0.40 | 2 |
| Accuracy |  |  | 0.80 | 15 |
| Validation Test (SUV features only) |  |  |  |  |
| Class | Precision | Recall | F1-Score | Support |
| Inflammation | 0.67 | 0.67 | 0.67 | 13 |
| Malignancy | 0.33 | 1.00 | 0.50 | 2 |
| Accuracy |  |  | 0.67 | 15 |

In contrast, the SUV-only model reached a lower overall accuracy of 0.67, with 8 true negatives, 2 true positives, 4 false positives, and no false negatives. Malignant lesions were always correctly identified (recall = 1.00), but this came at the cost of low precision (0.33), as several inflammatory lesions were misclassified as malignant. The classification of inflammatory lesions was less robust (precision = 0.67, recall = 0.67). Overall, these results indicate that the inclusion of parametric and derived radiomic features improves the balance between sensitivity and specificity. While the SUV-only approach maximized sensitivity for malignancy, it substantially reduced specificity, leading to overestimation of malignant findings. By contrast, the full feature set provided a more stable performance profile, with improved overall diagnostic reliability.

## IV. DISCUSSION

Current EANM/SNMMI guidelines recommend that absolute SUV values should be interpreted with caution in the context of infection and inflammation, as no validated cutoffs exist and uptake can overlap between benign and malignant processes. In line with this, our results confirm that SUV alone is insufficient to reliably discriminate between cancer and inflammation in oncologic imaging. This limitation reflects the fact that SUV integrates both metabolized and unmetabolized fractions of FDG without the ability to disentangle their respective contributions.

By contrast, our study demonstrates that SUV mean and first-order radiomic features, when evaluated alongside the corresponding Patlak-derived *K*_*i*_ and *V*_*d*_ values within the same VOI, allow a separation between malignant and inflammatory lesions. Parametric imaging explicitly decomposes the observed signal into metabolized uptake (*K*_*i*_, reflecting irreversible phosphorylation) and unmetabolized tracer (*V*_*d*_, reflecting tissue distribution volume). This framework provides a more physiologically meaningful interpretation of tracer behaviour than static SUV images alone.

Among the kinetic parameters, *V*_*d*_ is less directly interpretable than *K*_*i*_, but it can be considered a surrogate for the fraction of unmetabolized FDG retained within the tissue. Importantly, this parameter highlights differences in distribution patterns that static imaging cannot capture. In inflammatory lesions, the unmetabolized fraction tends to accumulate more peripherally, reflecting vascular delivery and limited metabolic trapping, whereas in malignant lesions FDG is rapidly phosphorylated and retained intracellularly, leading to higher *K*_*i*_ values and lower *V*_*d*_. This contrast between trapping and extracellular FDG distribution has been here proposed a reading approach to discriminate between malignant and inflammatory lesions.

Our findings diverge from previously published work, such as that of Skawran et al.[14], who reported only modest differences in Patlak-derived parameters between malignant and inflammatory lesions. One possible explanation is the difference in methodology. While previous analyses primarily compared global *K*_*i*_ or *V*_*d*_ values, we specifically interrogated the contribution of metabolized and unmetabolized tracer within SUV-defined VOIs (40% of SUVmax threshold). In this way, our approach acknowledges that SUV already integrates both components but lacks discriminatory power, whereas Patlak analysis allows us to quantify and map their distinct contributions within the same lesion volume. This difference in perspective likely underpins the separation observed in our dataset.

Taken together, these results suggest that short-dynamic Patlak imaging can provide a practical and physiologically grounded solution to the long-standing challenge of distinguishing cancer from inflammation. By moving beyond static SUV interpretation and explicitly assessing the balance between metabolized and unmetabolized FDG, we introduce a reproducible framework for clinical reading. While intuitive, this represents the first empirical demonstration of such tracer distribution patterns, offering a potential new standard for image interpretation in oncologic PET.

## V. CONCLUSION

This study demonstrates that short-dynamic Patlak FDG PET/CT, combined with rigorous preprocessing and machine learning, can enhance the differentiation of malignant from inflammatory lesions beyond static SUV analysis. By explicitly separating metabolized (*K*_*i*_) and unmetabolized (*V*_*d*_) tracer components, parametric imaging provides physiologically grounded insights that improve lesion characterization.

Our results show that radiomic features derived from *K*_*i*_/ *V*_*d*_ ratios and their distributions contribute significant predictive value, particularly when integrated into non-linear models. Addressing class imbalance with synthetic data further improved model robustness, yielding a clinically relevant balance between sensitivity and specificity. Importantly, this framework reduces the risk of both false positives and false negatives, underscoring its potential as a decision-support tool in equivocal cases.

Nonetheless, the present study is limited by its relatively small and imbalanced dataset, which may restrict generalizability. While synthetic oversampling mitigated some of these challenges, the results should be interpreted with caution. Future studies with larger, multi-centre cohorts are needed to refine model training, improve sensitivity for malignancy detection, and establish more robust and generalizable decision thresholds. Such validation efforts will be crucial to determine whether this approach can consistently support clinical differentiation of inflammation from cancer.

In summary, short-dynamic Patlak imaging, coupled with radiomics and machine learning, offers a reproducible and scalable strategy to overcome the long-standing challenge of distinguishing cancer from inflammation in FDG PET/CT. Prospective validation in larger and more diverse patient cohorts will be essential to confirm its clinical utility and facilitate translation into routine practice.

## AUTHOR APPROVAL

All authors have seen and approved the manuscript.

## ETHICS APPROVAL

This publication involved a prospective observational study. The data collected were fully anonymized and their use were approved by the Ethics Committee of IRCCS Humanitas Research Hospital (Approval ID: NUNC-001-2024, Prot. No. 351/24), in accordance with the principles of the Declaration of Helsinki.

## INFORMED CONSENT

Informed consent was obtained at the time of acquisition.

## DATA AVAILABILITY STATEMENT

The PET/CT imaging data used for this study are not publicly available due to patient privacy considerations.

