## Supplementary Materials for "Discriminating Inflammation from Malignancy with Short-Dynamic Patlak Parametric 18F-FDG PET/CT"

### Exploratory Data Analysis

**Table S1 –** Descriptive statistics (median and interquartile range) of SUV, $K_i$, and $V_d$ features for malignant and inflammatory lesions.

| **=== SUV ===** | **Malignant Lesion** | | **Inflammatory Lesion** | |
|---|---|---|---|---|
| **Feature** | **Median** | **IQR** | **Median** | **IQR** |
| Mean | 5.17 | 4.57 | 2.70 | 0.87 |
| Maximum | 8.68 | 6.88 | 4.40 | 2.05 |
| Variance | 1.67 | 2.59 | 0.44 | 0.30 |
| Skewness | 0.73 | 0.37 | 0.74 | 0.51 |
| Kurtosis | 2.62 | 0.80 | 2.76 | 0.74 |
| Entropy | 3.64 | 0.21 | 3.59 | 0.25 |
| **=== $K_i$ ===** | **Malignant Lesion** | | **Inflammatory Lesion** | |
| **Feature** | **Median** | **IQR** | **Median** | **IQR** |
| Mean | 1.61 | 1.90 | 0.64 | 0.41 |
| Maximum | 3.31 | 3.28 | 1.60 | 0.84 |
| Variance | 0.00 | 0.01 | 0.00 | 0.00 |
| Skewness | 0.44 | 0.32 | 0.48 | 0.37 |
| Kurtosis | 2.51 | 0.58 | 3.04 | 0.80 |
| Entropy | 3.72 | 0.16 | 3.54 | 0.20 |
| **=== $V_d$ ===** | **Malignant Lesion** | | **Inflammatory Lesion** | |
| **Feature** | **Median** | **IQR** | **Median** | **IQR** |
| Mean | 30.68 | 35.23 | 37.46 | 26.29 |
| Maximum | 94.63 | 71.84 | 123.94 | 62.36 |
| Variance | 338.67 | 717.74 | 571.70 | 731.14 |
| Skewness | 0.46 | 1.19 | 0.39 | 0.57 |
| Kurtosis | 2.91 | 1.17 | 2.73 | 0.53 |
| Entropy | 3.43 | 1.04 | 3.58 | 0.20 |

**Table S1 –** Normality (Shapiro-Wilk) and differences (Mann-Whitney U) tests divided by classes. The Shapiro-Wilk test revealed that the majority of features in both the malignant and inflammatory groups did not follow a normal distribution. The Mann-Whitney U test, used to assess differences between the two lesion types, showed that 10 out of 18 features had a statistically significant difference ($p<0.05$). This indicates that many of these radiomic features, when analysed individually, hold potential for distinguishing between lesion types.

| **=== SUV ===** | **Shapiro-Wilk p-value** | | **Independency** | |
|---|---|---|---|---|
| **Feature** | **Malignant** | **Inflammatory** | **MWU** | **Significant** |
| Mean | 0.0110 | 0.1014 | 0.0000 | Yes |
| Maximum | 0.0010 | 0.0222 | 0.0000 | Yes |
| Variance | 0.0000 | 0.0001 | 0.0000 | Yes |
| Skewness | 0.2655 | 0.1264 | 0.8577 | No |
| Kurtosis | 0.0001 | 0.0000 | 0.0576 | No |
| Entropy | 0.0082 | 0.0001 | 0.0916 | No |
| **=== $K_i$ ===** | **Shapiro-Wilk p-value** | | **Independency** | |
| **Feature** | **Malignant** | **Inflammatory** | **MWU** | **Significant** |
| Mean | 0.0007 | 0.0011 | 0.0000 | Yes |
| Maximum | 0.0032 | 0.0020 | 0.0000 | Yes |
| Variance | 0.0000 | 0.0001 | 0.0000 | Yes |
| Skewness | 0.4533 | 0.0001 | 0.1380 | No |
| Kurtosis | 0.0004 | 0.0000 | 0.0017 | Yes |
| Entropy | 0.0000 | 0.0016 | 0.0002 | Yes |
| **=== $V_d$ ===** | **Shapiro-Wilk p-value** | | **Independency** | |
| **Feature** | **Malignant** | **Inflammatory** | **MWU** | **Significant** |
| Mean | 0.0085 | 0.0547 | 0.0425 | Yes |
| Maximum | 0.0000 | 0.2030 | 0.2216 | No |
| Variance | 0.0000 | 0.0000 | 0.3877 | No |
| Skewness | 0.0046 | 0.0965 | 0.0981 | No |
| Kurtosis | 0.0000 | 0.0001 | 0.1296 | No |
| Entropy | 0.0000 | 0.0000 | 0.0145 | Yes |

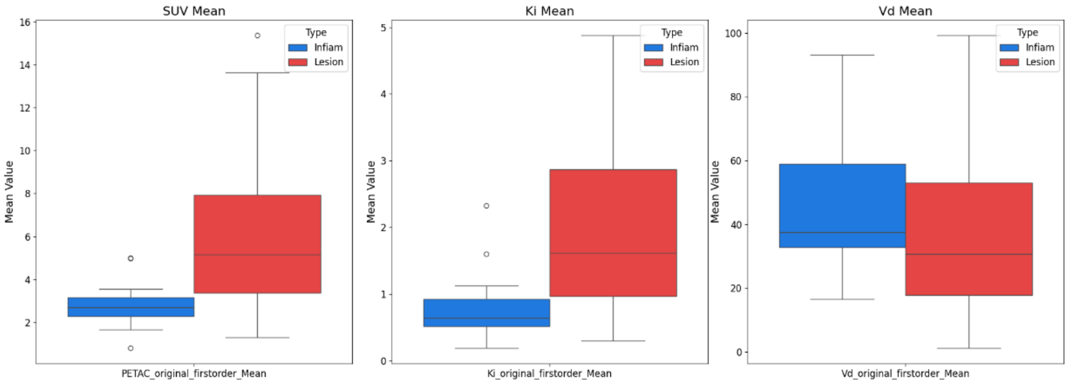


**Figure 1S –** Boxplots of SUV, Ki, and Vd for malignant and inflammatory lesions.

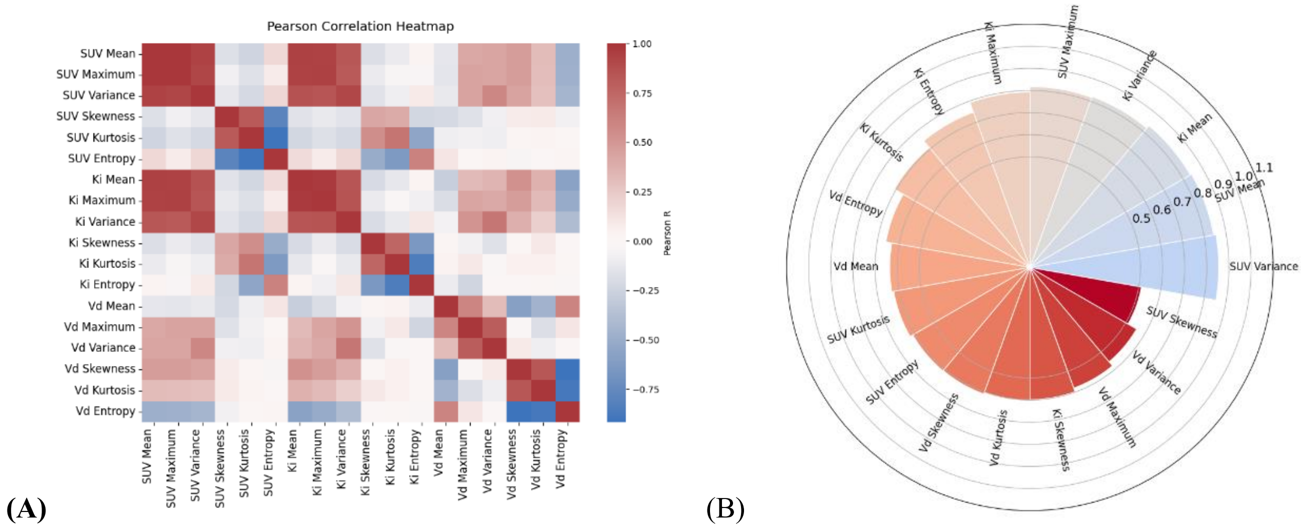


**Figure 2S –** Summary of the exploratory data and linear separability analysis. **(A) Pearson Correlation Heatmap.** A heatmap illustrating the Pearson correlation matrix of the 18 radiomic features. The colour intensity and value for each cell represent the correlation coefficient (r), with colours ranging from blue (strong negative correlation) to red (strong positive correlation). **(B) Single-Feature ROC-AUC Radial Bar Chart.** This radial bar chart illustrates the discriminatory power of each individual radiomic feature. The values on the radial axis represent the AUC, ranging from 0.5 (random classification) to 1.0 (perfect classification). Each bar corresponds to a feature, and its length indicates its AUC score.

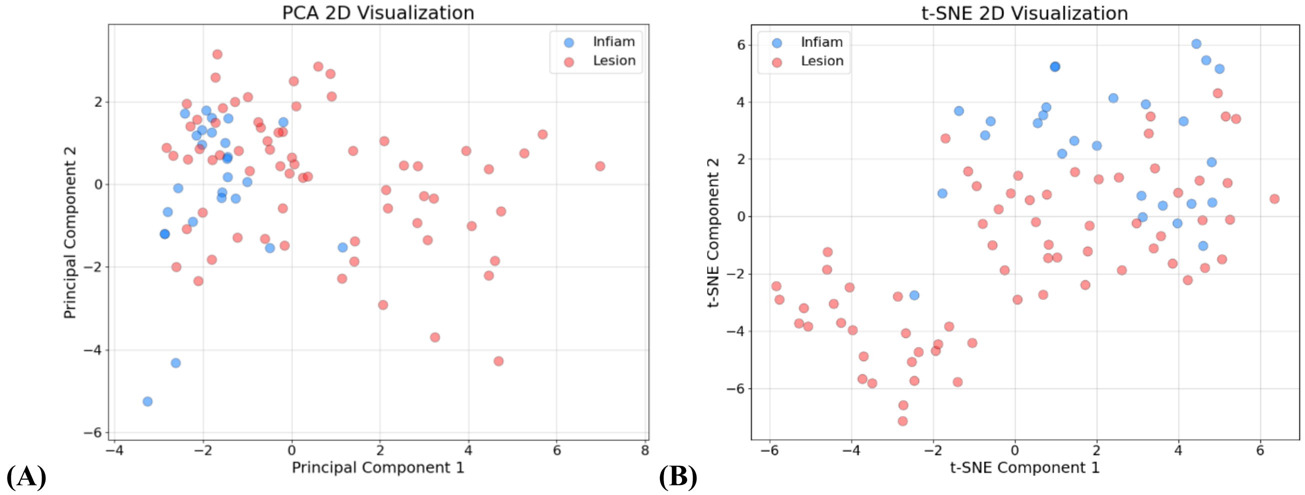


**Figure 3S** – Summary of the exploratory data and linear separability analysis. **(A) PCA 2D Visualization.** Scatter plot showing the distribution of samples in a two-dimensional feature space, following dimensionality reduction via PCA. The plot visually demonstrates significant overlap between the two classes, confirming that the data is not linearly separable. **(B) t-SNE 2D Visualization.** A scatter plot of the two-dimensional representation of the high-dimensional data, generated using t-SNE after data standardization. Similar to the PCA plot, this visualization reveals that the malignant and inflammatory lesions are not clearly separated into distinct clusters.
